# A data-driven epidemiological model to explain the Covid-19 pandemic in multiple countries and help in choosing mitigation strategies

**DOI:** 10.1101/2020.08.15.20175588

**Authors:** M. J. Beira, A. Kumar, L. Perfeito, J. Gonçalves-Sá, P. J. Sebastião

## Abstract

Accurate models are fundamental to understand the dynamics of the COVID-19 pandemic and to evaluate different mitigation strategies. Here, we present a multi-compartmental model that fits the epidemiological data for eleven countries, despite the reduced number of fitting parameters. This model consistently explains the data for the daily infected, recovered, and dead over the first six months of the pandemic. The good quality of the fits makes it possible to explore different scenarios and evaluate the impact of both individual and collective behaviors and government-level decisions to mitigate the epidemic. We identify robust alternatives to lockdown, such as self-protection measures, and massive testing. Furthermore, communication and risk perception are fundamental to modulate the success of different strategies. The fitting/simulation tool is publicly available for use and test of other models, allowing for comparisons between different underlying assumptions, mitigation measures, and policy recommendations.

---

We are currently fighting to control a newly emerged infectious disease (COVID-19) caused by severe acute respiratory syndrome coronavirus 2 (SARS-CoV-2). As we write, COVID-19 has expanded to touch nearly every country in the globe, infecting more than 19 million people worldwide, and claiming more than 700,000 lives.^1^ This happened despite what might be considered the largest lockdown in history, with authorities implementing several preventive measures from social distancing to isolating entire countries. These restrictions have been instrumental in reducing the impact of the pandemic, but most decision-makers acknowledge that it will be necessary to further loosen the confinement measures even in the absence of any effective vaccine or treatment. In the interest of economic recovery, several countries are deploying different measures such as conditional movements and contact tracing apps. Others have already re-opened schools and restaurants or are preparing to do so.

For as long as we are limited to non-pharmaceutical interventions (NPIs), models that can accurately describe the dynamics of the disease and allow for *in silico* testing of different scenarios are of utmost importance for scientists, health authorities and decision-makers in general. In the past, such models have been proven very valuable, including in coronavirus outbreaks.^2,3^ The retrospective analysis of the models along with the fact that we can already see the result of different strategies, from Japan’s decision not to lockdown to Iceland’s massive testing, offers the unique opportunity to test whether a single model can incorporate all these differences, in close to real-time. In the case of the current pandemic, a variety of models have been published or publicly released, which can be broadly organized into three groups: 1) statistical models, which can allow researchers to broadly forecast the progression of the disease and how many people are likely to die; 2) extended SIR-type mathematical models, which can potentially answer questions about the effect of the behavior of each individual in a compartmentalized population and the effect of general preventive measures (mask-wearing, social distancing, quarantine, and lockdown) on the disease spread, and 3) data and computationally intensive agent-based networks, which use individual-level information on mobility, socio-economic conditions, etc., to make very complex descriptions of interactions.^4–13^ Some of these models have shown great differences in their predictions and the reported numbers and there is little consistency among them.^14^ Several possible reasons can explain such variability. First, COVID-19 has affected almost every country and the evolution of the disease varies widely among them (Fig.1). Second, the behavior of COVID-19 disease is unique in many ways (ex. common asymptomatic spreading that forced the first-ever mass-level lock-down). Third, unlike past pandemics, most predictions are happening in close to real-time, dealing with inaccuracy in the reported data, large differences in testing levels, and individual behaviors. Fourth, many models assume a *closed population*, ignoring demographic changes or travel influx/outflux. This was never the case in most countries (including in Europe) as the borders were never closed, and a fraction of the population was always moving in and out. Hence, most of the existing models, let alone the basic SIR model, remain too coarse-grained, not including crucial factors such as the impact(s) of quarantine, pre-symptomatic (silent) spreading, and any variation in the susceptible/infected population, when projecting the behavior of the disease.^15^

**Fig. 1.**
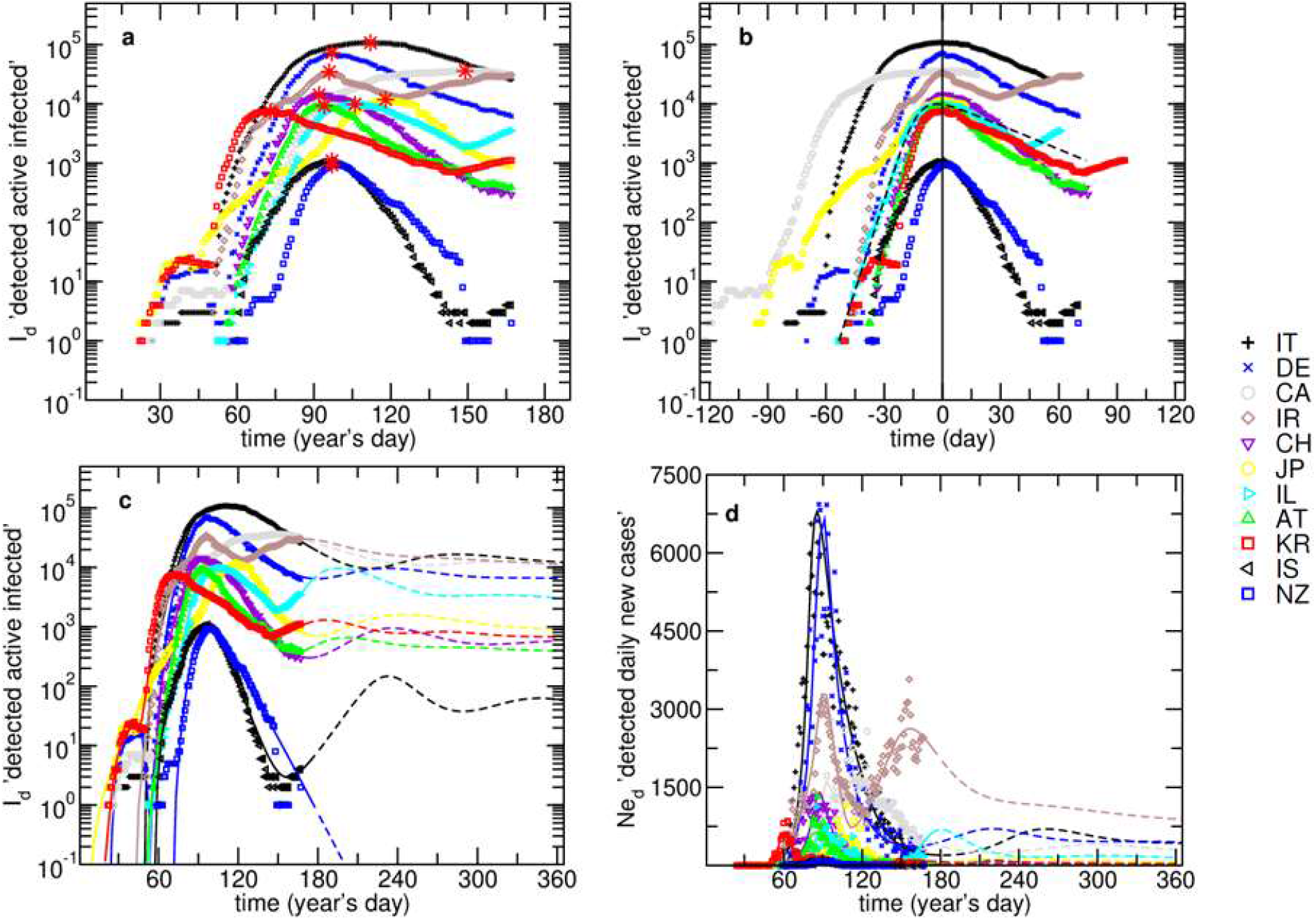
Time evolutions of detected active infected, ***I**_d_*, and daily new cases, ***Ne_d_***, for **IT, DE, CA, IR, CH, JP, IL, AT, KR, IS** and **NZ**. (**a**) – ***I_d_*** evolution (red asterisks mark the maxima); (**b**) – ***I_d_*** centered around the peak (black dashed line shows the SIR model fitting curve for **IL**); (**c**) – ***I_d_*** with model fitting curves; (**d**) – ***Ne_d_*** with model fitting curves. The presented curves are partially dashed in order to separate the data fits from the projections. The model fitting is explained in the text.

Conversely, it can be argued that more complex models might mask important features of the disease and become too distant from the underlying biology and epidemiology of infectious agents.^7,16^ Moreover, overly complex models are often too dependent on the parameters and data used, losing generality. However, they can project the future dynamics of the epidemic, the effectiveness of mitigation measures, and even the re-appearance of the virus.

In this study, we integrate epidemiological data from 11 countries and develop a nine-compartimental model, PSEIRD(S), to consistently fit all data. The method followed additionally allows to focus on three fundamental problems: 1) additional outbreaks, 2) the role of individual behavioral changes (such as mask wearing, self-quarantine) and 3) the role of public policies (such as increased testing, contact tracing or lockdowns) in mitigating such outbreaks.

## Results

Figure 1a shows the daily evolution of the number of detected actively infected people, *I_d_*, since the beginning of this year and in Fig. 1b this evolution is centered around the date it peaked. The maxima (Fig 1a, red asterisks) appeared at different times and the number of cases varies over orders of magnitude, for the presented countries. However, as it is often the case with infectious diseases, the behavior of the curves is similar (Fig 1b), with an initial growth that depends on the presence of infected people in a susceptible community, which evolves depending on confinement measures, followed by a decrease influenced by de-containment measures, recoveries, and deaths. It is important to note that the simplest SIR – Susceptible, Recovered and Infected – model can fit the first-wave data satisfactorily (Fig. 1b, black dashed line) but: a) does not explicitly represent the actual behavior of people (ex. travelling, quarantine and wearing masks), b) cannot predict second outbursts, or waves, unless we assume re-infection, c) considers that all individuals initially in the susceptible compartment, ***S0***, will eventually be infected (unless the recovery rate is sufficiently larger than the transmission rate). If we assume that the susceptible group is equal to the entire population of a country, we may reach a conclusion that around 70% of the entire population will be infected, leading to the collapse of any health care system (see *Normalized SIR model fit for Austria* subsection in the supplementary materials).^17^

The PSEIRD(S) model (Fig. 2) builds on previous extensions of the SIR model and expands them to better represent the current pandemic (and possibly any pandemic disease), by allowing for open populations and discrimination of flows among compartments.^3,6,15,18^

**Fig. 2.**
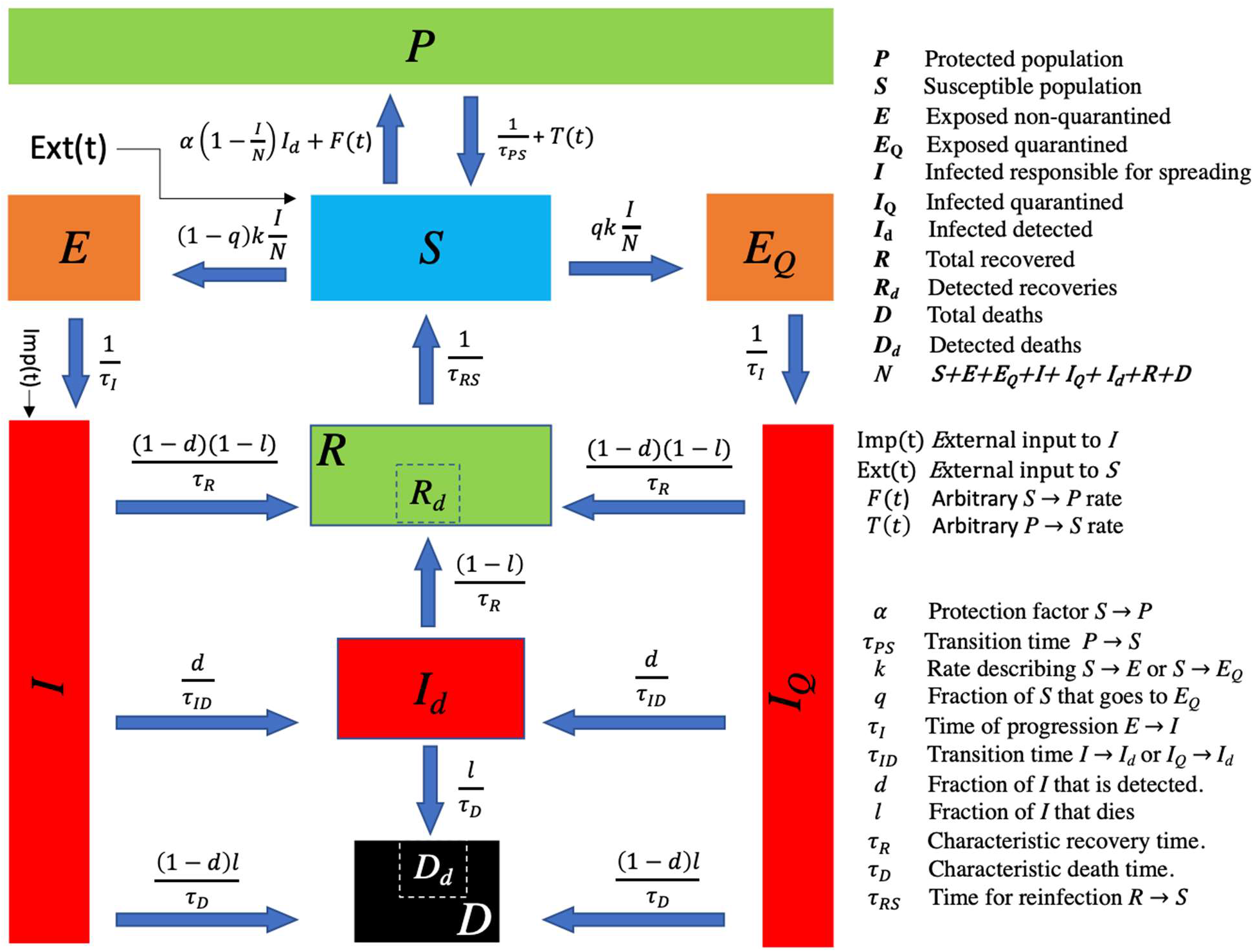
Schematic representation of the nine-compartment PSEIRD(S) model. The parameters are summarized in the figure and described in more detail in Supplementary Table S1.

One important feature of the PSEIRD(S) model is the existence of a protected compartment ***P***, that represents a part of the population that does not participate in the infectious process, for example, because of geopraphical reasons, self-protection measures or because they are just lucky enough never to come in contact with an infectious individual. In this way, PSEIDR(S) is more general and suitable to describe the data available for the current pandemic and, we believe that, for any infectious disease. A complete description of the PSEIRD(S) model is presented in the *Model* subsection of the Supplementary Text.

Tab. 1 shows the fitted parameters and initial sizes of the susceptible and infected compartments (***S*** and ***I***) for each country. The initial values for the remaining compartments can be found in the Supplementary Fits Reports.

**Table 1:**
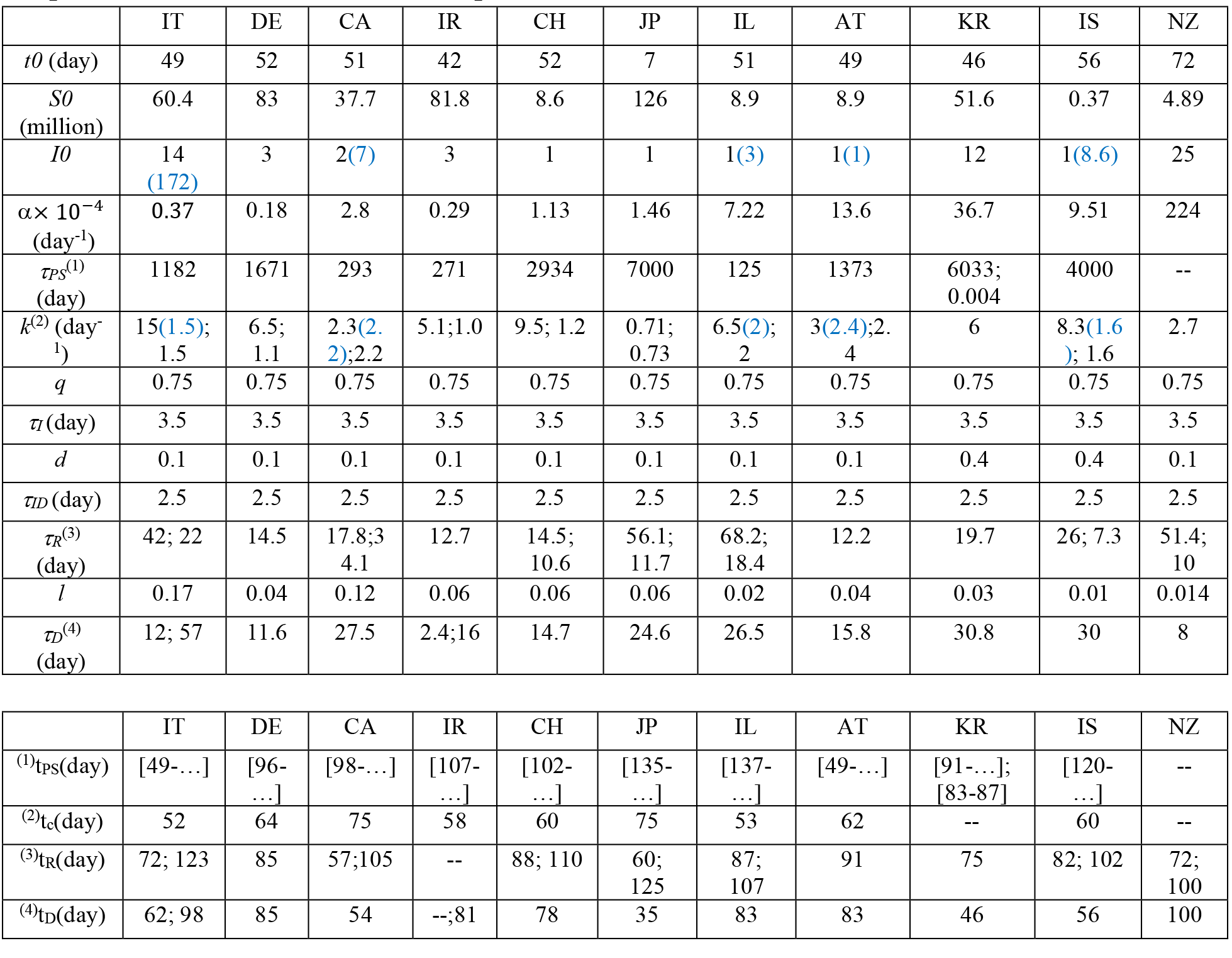
Model fitting parameters obtained for the studied countries. Initial values for the compartments ***S*** and ***I*** are also provided. The initial values for the remaining compartments and the parameters uncertainties can be found in the Supplementary Fits Reports and in Fig. 4, respectively. The multiple values assumed by one parameter are separated by a semicolon. The moment that sets the change is described by the delay associated with that parameter (***k*** relates to ***t_c_***; ***τ_R_*** relates to ***t_R_***; ***τ_D_*** relates to ***t_D_*** and ***τ_PS_*** relates to ***t_PS_***, as explained in Supplementary Table S1). The highlighted numbers were obtained for one possible fit where ***k_1_*** was fixed and equal to ***k_2_*** and ***I0*** was optimized to accommodate this change. For the cases where this alternative fit was not possible, no additional values are presented.

The model-fitting results for ***Id*** and ***Ned*** are shown in Figs 1c and 1d, respectively. The complete set of fitting compartments (***Ne_d_, NT*_d_, *D*_d_, *R*_d_** and ***I*_d_**) is shown in Fig. 3 and Supplementary Figure S2. In Fig. 3 are presented the results for **DE, IL** and **KR** that applied different mitigation strategies and have different dynamics. It can be observed that the model anticipates the appearance of additional waves in several countries. This happens despite the scenarios being different for each country and described by different values of the model parameters (Tab. 1). The dashed lines in the figures represent the projections made until the end of the first year, assuming that the model parameters remain constant over this period.

**Fig. 3.**
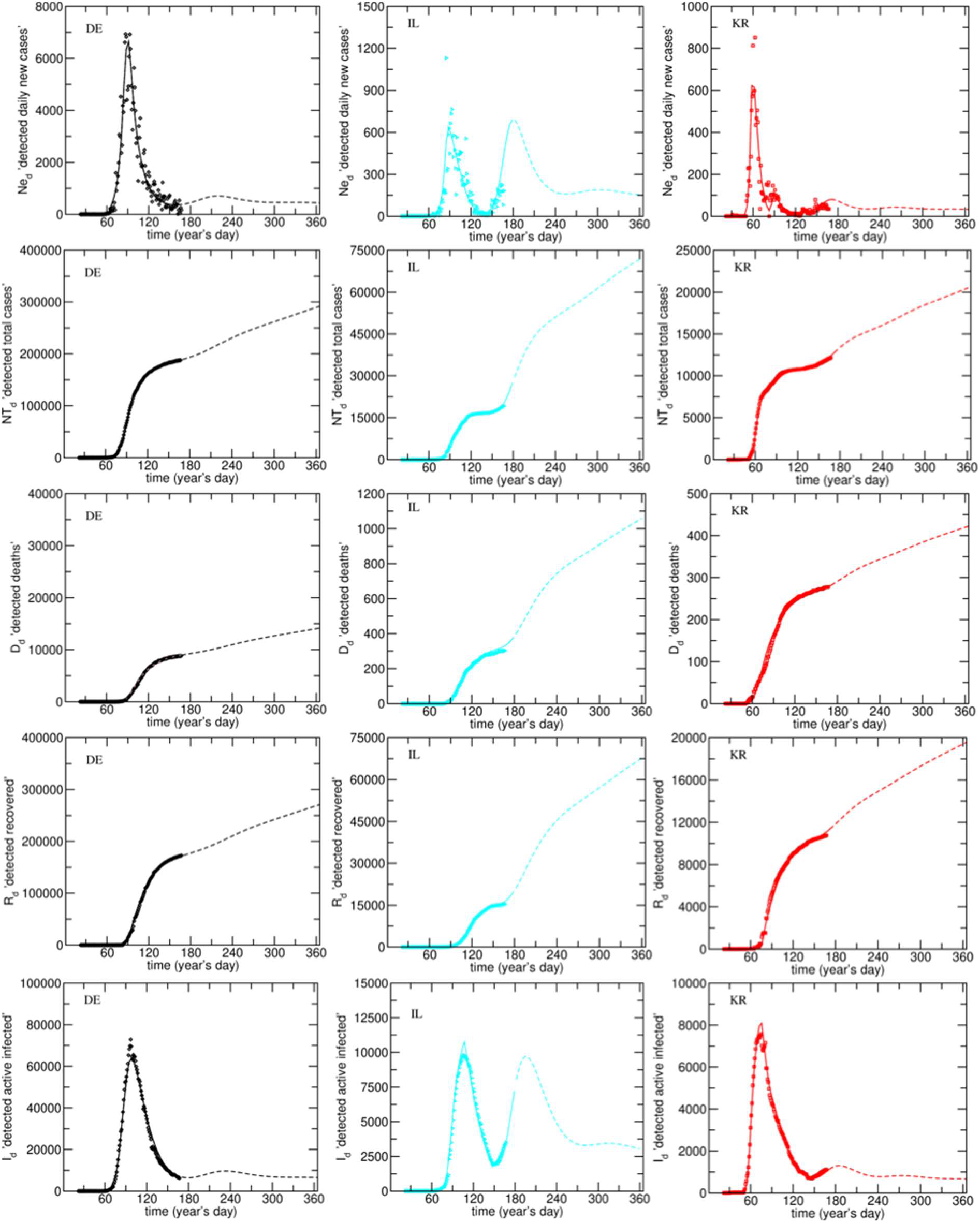
Model fits obtained for **DE, IL**, and **KR** (remaining countries in Supplementary Figure S2). Countries in columns, compartments in rows. The dashed lines are the projections as explained in the text.

The first small epidemic outburst, observed for **DE, CA**, and **KR**, was separately analyzed and the obtained results can be found in the Supplementary Fit Reports. This is related to the fact that the initial spreading of the disease can hardly be described by the same parameters as those used for the main epidemic outburst. However, it was possible to make these separate fits compatible by setting the initial conditions for the second outburst based on the compartmental variable obtained from the model fitting to the first.

The fact that the model is able to accurately explain the data for such an extensive selection of countries and of compartments demonstrates its robustness and potential and allows for extensive analysis. The fits were simultaneously obtained in a manageable way mainly due to the use of a powerful fitting tool. This open-access platform is available for use and test of other models.^20^ In the Supplementary Fit Reports, it is also possible to access the simulation of the remaining unknown compartments (***P, S, E, E_Q_, I, I*_Q_, *R, D***, and ***NT***) for all the analyzed countries. It is important to note that the simulated compartments were subject to the same reporting delays found for the fitted compartments. This produces a simulation that provides the worst-case scenario for ***I*** and ***I***_Q_, ***I*** being the compartment that, along with ***S***, enables the spreading of the disease.

A particularly interesting difference among the countries was in the confinement policies and the percentage of the population that remained in isolation. As individuals become less disciplined or the government lifts some strict confinement measures, the model projects the existence of additional peaks. These peaks result from the transition of people from compartment ***P*** to ***S*** (***PS***-leakage), which were observed for all countries, except **NZ**. In the model, this transition was included either by a continuous ***PS**-*leakage in time (when the amplitude of the secondary infection is not yet known) or by a ***PS***-leakage acting within a given time range that can only be estimated when the maximum of the second outburst has been reached.

The ***Ne_d_*** time series provides relevant information regarding different types of ***PS***-leakages. When there is no ***PS***-leakage and the detection characteristics are maintained, the distribution observed for the ***Ne_d_*** will monotonously decrease to zero. If a tail or additional peaks are observed, there is a clear indication of a ***PS***-leakage. ***Ne_d_*** data analysis/simulations that disregard a possible change in the number of susceptible people with time will, in most cases, not be able to explain the reported data available for this compartment and may lead towards erroneous conclusions regarding the end of the epidemic.^22^

In the case of **KR**, it was possible to distinguish two different outbursts in the ***Ne_d_*** evolution (Fig. 3) and an additional third, whose amplitude is still unknown. Only in the case of **IT** and **AT** it was possible to fit a continuous ***PS***-leakage, acting from the beginning of the epidemic. In the remaining cases, the ***PS***-leakage was more significant, and its onset was optimized by the fit. This may be due to a later or less abrupt confinement/deconfinement transition in **IT** and **AT** or to a reduced number of remaining infected people at the beginning of deconfinement, particularly for **AT**.

### Model fitting parameters

The uncertainties of the parameters presented in Tab. 1 can be seen in Fig. 4 and were estimated by assessing the sensitivity of the global least-squares minimum to the parameters when they are simultaneously set free to optimize. As there are several compartments characterized by values with different orders of magnitude (compare, for example, the detected dead with the total number of detected cases), the χ^2^ was normalized to 1 for each compartments in order to extract the uncertainties of the fitting parameters.

**Fig. 4.**
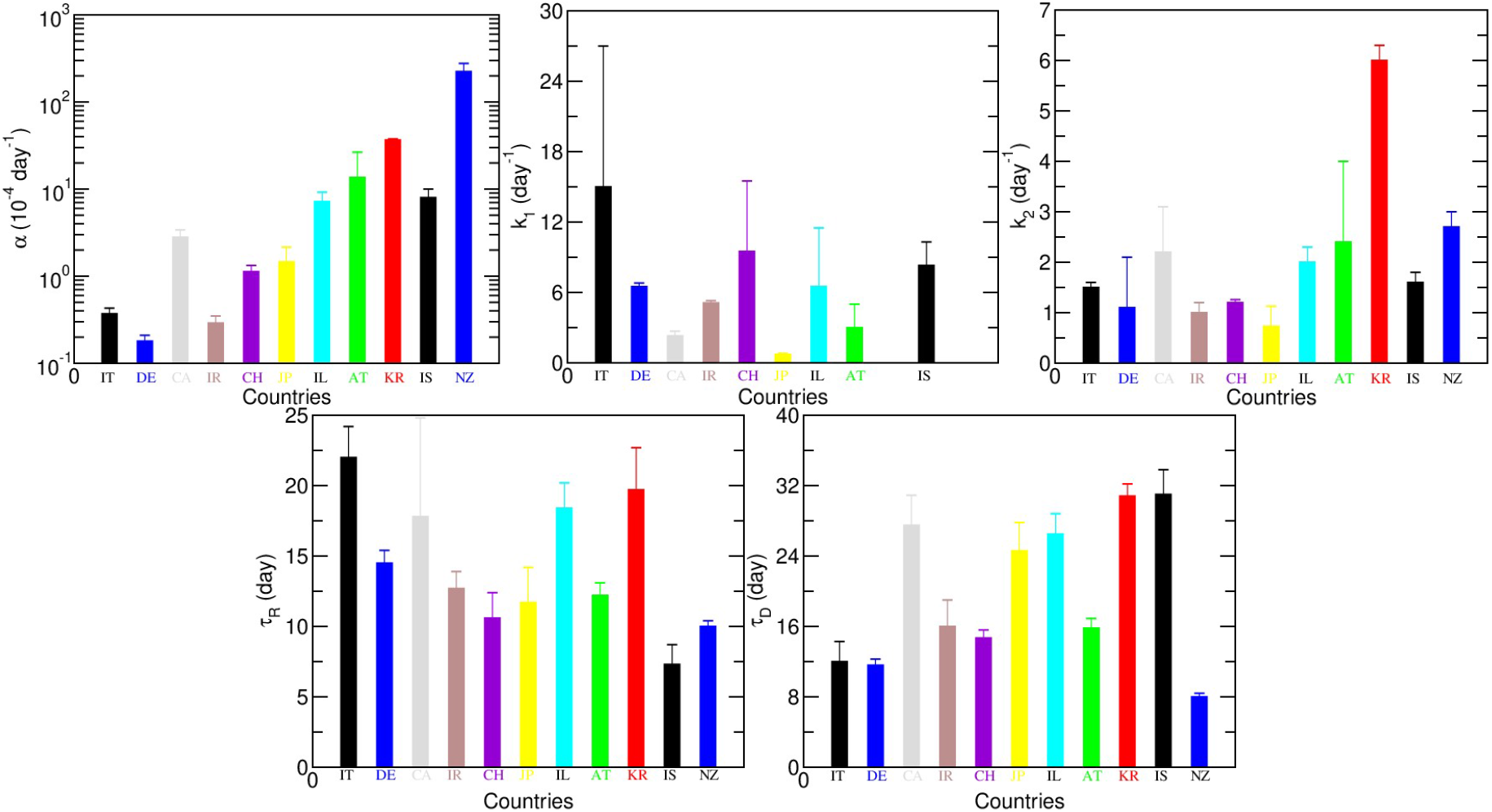
Fitting parameters values and uncertainties obtained for each of the studied countries. Uncertainties were determined as explained in the text.

As can be observed in Fig. 4, α is a well-estimated parameter although, unsurprisingly, presents vastly different values for the studied countries. This happens because of the difference in population sizes, on the one hand, and because of the protective measures applied by each country, on the other.

As there are not enough data or direct means to estimate some of the model parameters such as ***q***, *τI*, ***d*** and *τI**D***, rather than optimizing these parameters, we fixed them to reasonable values. For the parameter ***q***, it is rational to expect that 75% of the people comply with the isolation/quarantine measures even in the extreme lockdown situation.*τI* was assumed to be the incubation time of the disease (about 5 days) minus 1–2 days during which, even though asymptomatic, infected people can transmit the disease^*23*^. Both ***q*** and *τI* are correlated parameters although the margin within which *τI* can be varied is rather small.

The realistic values of ***d*** will only be known when more serologic test results are obtained. So far, some studies have suggested that between 14% to 20% of the population may have been infected, which provides detection percentages of the order of 2–3% for most countries^*24*^. This may not be realistic because of the heterogeneity associated with the transmission of the disease within a city or a country and because the sample choice for these tests needs to be very randomly selected. The value of ***d*** was set to 10% except for the cases of **KR** and **IS**, which could not be fitted with such a small value (most probably due to contact tracing in the case of **KR** and extensive testing in the case of Iceland, which has tested almost half its population). The parameter *τI**D*** represents the time it takes to detect an infectious person and, excluding the delays caused by the processing of the results, we assumed that the process could take up to 2–3 days from infectiousness. As ***d*** and τ_IPS_ are also correlated to each other, it is quite difficult to attribute significantly different values of *τI**D*** for a given value of ***d***.

Except for **KR** and **NZ**, the data required the use of two different values of ***k***. This parameter, which contains the number of daily contacts times the probability of a contact generating an infection, is assumed to vary discretely with time instead of continuously. It could be argued that there would have to be more than two values of ***k***, namely one for each pack of restriction/containment or loosening measures applied. However, trying to add more variables would over parametrize the problem. In fact, as can be observed in Fig. 4, ***k**_1_* is already associated with an uncertainty much larger than the remaining parameters and could not even be estimated for the case of **KR** and **NZ**. This is understandable, since ***k**_1_* is estimated using a much smaller number of points, since only the portion of the graph where t < t_c_ is affected. The value of this parameter is only perceived if we observe the detected active infected cases in a logarithmic scale (as shown in Fig. 1c or the Supplementary Fits Reports). ***k**_2_* is easier to estimate, as evidenced in Fig. 4, but some striking differences, for example, between the values obtained for **JP** and **KR** are immediately apparent. In South Korea, this value remained high most probably as a result of massive contact tracing (people feel secure to continue their normal lives) while in Japan it was so low (due to massive mask-wearing and social distancing) that the disease did not spread as much as it could, even without the application of very strict confinement measures.

The existence of two ***k***s may also be a consequence of the misestimation of the initial value of the infected non-isolated cases. In fact, it is possible (for some of the countries) to produce good fits using a single value of ***k*** and letting ***I0*** as a free model fitting parameter (see values obtained for the alternative fit – in blue in Tab. 1). As the deconvolution of these two processes is not easy, we have provided the two contrasting alternatives.

*τ**R*** and *τ**D*** are parameters that can be well determined using properly reported recovered and dead data. Moreover, having the data sets for the recovered and dead is essential for the correct estimation of *τ**R*** and *τ**D***. This happens because the process of reporting data, especially for the recovered, may be subject to delays. These delays (parameters ***tR*** and ***tD*** in Tab. 1, explained in Supplementary Table S1) cannot be extracted using the active infected data and, unaccounted for, lead to an overestimation of the characteristic recovery/death time. If the reporting process for deaths and recoveries is systematic, the delays can be extracted from the fit and provide a good estimation both for *τ**R*** and *τ**D***. Examples of poor reporting of the recovered data are Norway and Portugal. Both these countries started almost by not reporting recovered cases at all and finished by having more than 10000 recovered cases in one day, which generated large discontinuities in the evolution and makes it impossible to determine the characteristic recovery time^19^.

The characteristic death time is generally relatively easier to obtain and is subject to less delays, although, in the case of Iran and Italy, two values of *τ**D*** had to be used in order to provide a good numerical fit, one of them being nonsensical in terms of the reality of the disease. This may have happened either because the method of reporting was changed or because it was inconsistent among different regions of these two big countries. *τ**R*** is much more susceptible to reporting inconsistencies, and therefore, the use of two values was more frequently needed. This explains why the uncertainties of this parameter are larger when compared to *τ**D*** (Fig. 4).

To discriminate between the dead and recovered compartments, the value of ***l*** (fraction of people that die) was calculated by dividing the total number of dead by the sum of dead and recovered, taken at day 167 (last data point).

### Simulations

In the case of **IL-** Fig. 3, the data suggest that around day 140 (May 19th) a leakage is observed, which leads to the increase both ***Ned*** and ***Id***. This is consistent with the dates at which Israel lifted their more strict measures: for example, schools started to re-open from early May to May 17^th^. If the leakage which is necessary to accommodate such an increment is applied indefinitely, future outbursts will be unavoidable as well as the endemic characteristic of the disease (at least for one year). In the case of **DE** and **KR**, if the current situation is maintained, secondary “waves” are likely of smaller amplitude, in comparison to the main outbreak. The secondary peaks can become as large as the first one, or even larger, if the leakage increases, as illustrated for **IL**.

However, many parameters can be acted on in order to prevent such outbursts, namely controlling the fraction of the population that goes into self-isolation after being exposed (***q***), the fraction of detection (***d***), and the time until which this leakage is active.

As mentioned, the projected curves in Figs. 1 and 3 were obtained assuming that none of the fitting parameters would change over one year. As this is unlikely, it is important to project and evaluate different scenarios. The results obtained from these simulations are presented in Fig. 5 and 6.

**Fig. 5.**
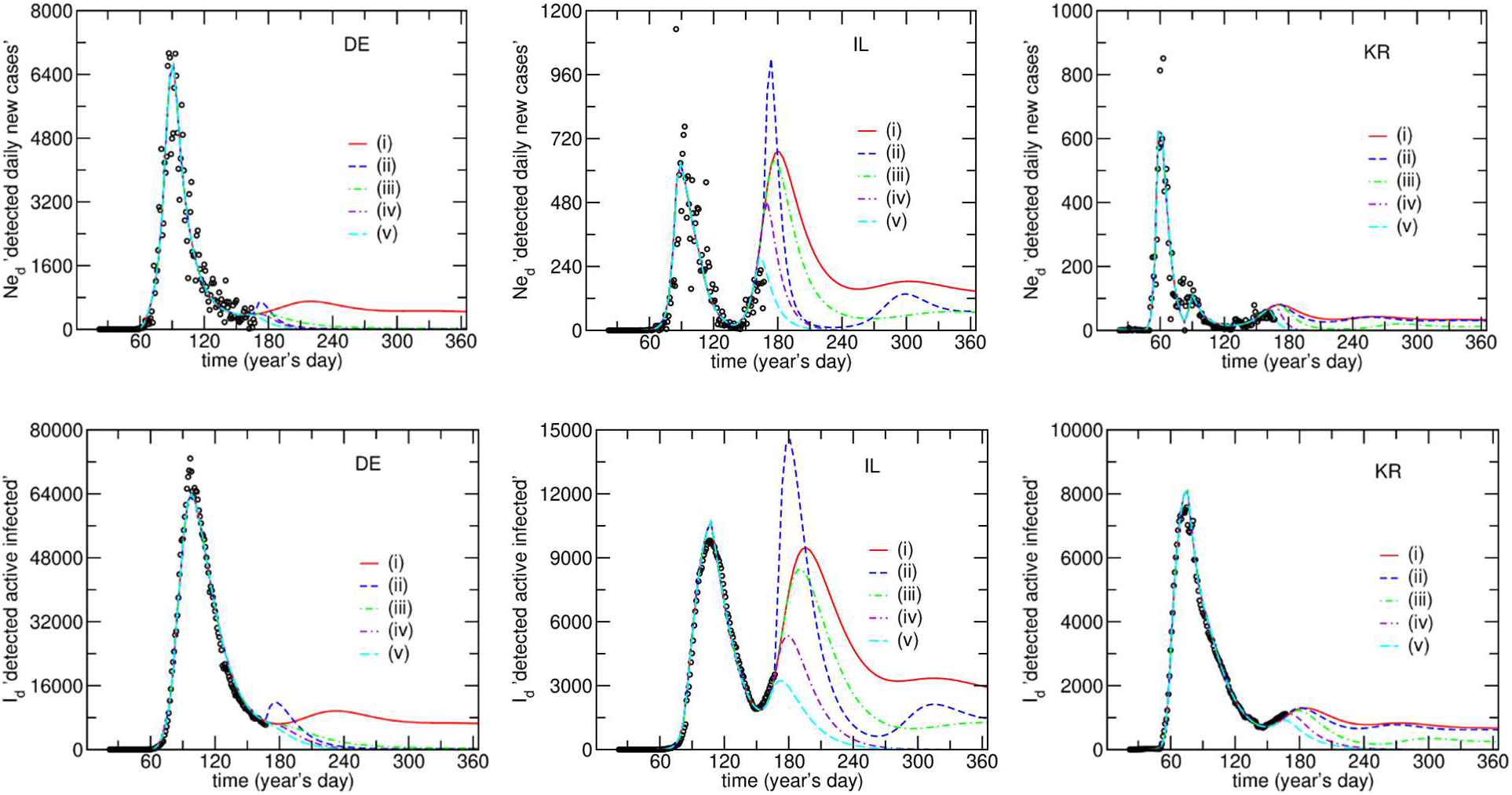
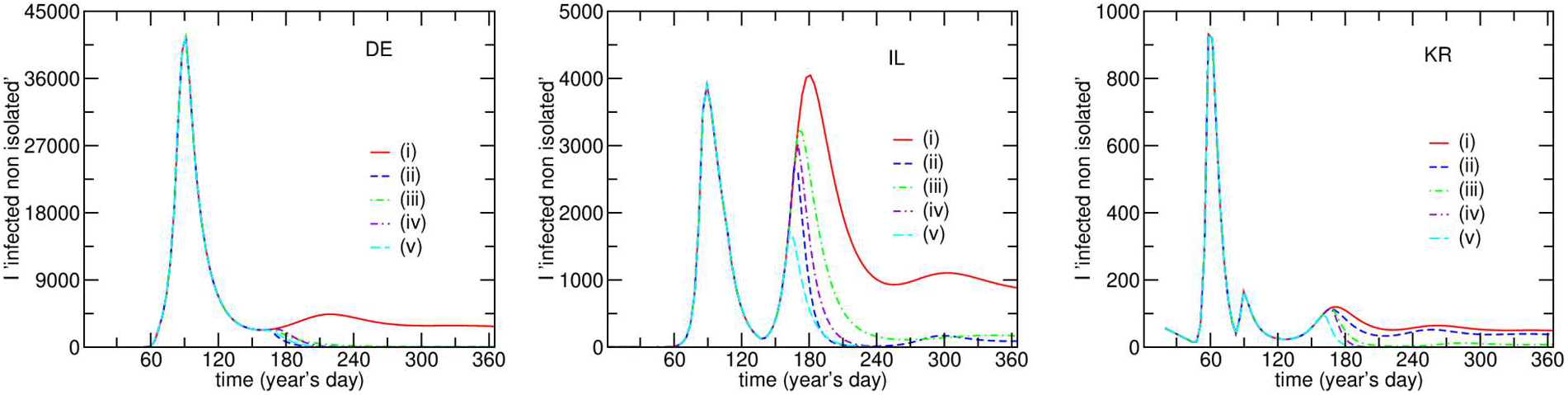
Model fits and simulation of possible scenarios for the ***Ne_d_**, **I_d_*** and ***I*** compartments based on the parameters obtained for the case of **DE, IL**, and **KR**. The model projects a second infection as a result of the ***PS***-flow with a rate obtained from the best fit and keeping all other parameters constant. (i) the rate of flow of people from ***P*** to ***S*** will continue until day 365 and nothing else changes. (ii) the rate of flow of people from ***P*** to ***S*** will continue until day 365 and ***d*** is increased to 0.5 after day 167. (iii) the rate of flow of people from ***P*** to ***S*** will continue until day 365 and ***q*** increases to 90% after day 167. (iv) the rate of flow of people from ***P*** to ***S*** stops at day 167. (v) the rate of flow of people from ***P*** to ***S*** stops on day 160.

Fig. 5 depicts the time evolution of ***Ne_d_**, **I_d_*** and ***I***. It is important to analyze all of these compartments in order to properly evaluate which parameter is more effective in reducing the amplitude of the second outburst.

The effect of increasing ***d*** to 50% (Fig. 5; scenario (ii)) is substantially positive, in view of the drop in ***I*** (responsible for spreading the disease), and despite a small initial increase in ***I*_d_** and ***Ne*_d_**, due to the increase in testing/detection ***d***. This is expected, since making more tests will initially lead to an increase in the detected population but reduce the effects produced by the ***PS***-leakage as the compartment ***I*** is depleted of people. For the countries whose ***d*** value was set to 0.4 (**KR** and **IS**), the increase of ***d*** to 0.5, naturally does not have the same impact observed for the remaining countries, whose initial ***d*** value was 0.1 (Fig. 5 and Supplementary Figure S3).

Interestingly, varying the percentage of the exposed population in self-isolation, ***q***, produces a much smaller effect than ***d*** and is probably the least realistic change made for this simulation (it assumes that 90% of the exposed population self-isolates (Fig. 5; scenario (iii)). This result is also important to understand the potential use of mobile applications to keep the disease from spreading since an application that works only by alerting potentially exposed people would mostly act on ***q***. In the case of a continuous leakage, it is clearly not the best parameter to act upon since it needs a very large percentage of warned and complying people, which is an unrealistic assumption in most countries.

If the leakage is stopped after 30 days (Fig. 5; scenario (iv)), it is possible to observe that the second peak is appreciably reduced and, although the effect of increasing *d* to 50% remains larger, stoping the leakage after day 167 seems to be a better option than increasing *q* to 90% (in the event of one having to choose between taking one option or the other). If the stopping of the leakage is anticipated seven days (Fig. 5; scenario (v)), however, it is clear that the second peak becomes much smaller than what was obtained for any of the other scenarios. Moreover, stopping the ***PS***-flow seems to be the only option to prevent the disease from becoming endemic in the population. The leakage from the ***P*** to the ***S*** compartment is the most important parameter to act upon if one wants to reduce the spreading of the disease, since it is applied to the large compartment ***P*** and, therefore, can feed the **S** compartment with a lot more individuals than the ones that tested positive so far for any country. The way to act on this leakage is to promote contact tracing mechanism, like the ones used in **KR**, and/or self-containment of the virus in a way where everybody acts as if they were infected (wear masks and follow social distancing), like in **JP**. The latter solution requires that the population develops self-awareness and discipline and is also an indirect way of acting on ***q*** without the need for lockdown.

The scenarios presented in Fig. 5 were also tested for the remaining countries except for **NZ**, since this country was not projected to have a ***PS***-leakage-based second outburst (Supplementary Figure S3).

As several of these parameters co-vary, additional simulations for **IL** are presented in Fig. 6 where it is easier to observe the cross-effect between ***q**, **d*** and the ***PS***-leakage duration.

**Fig. 6.**
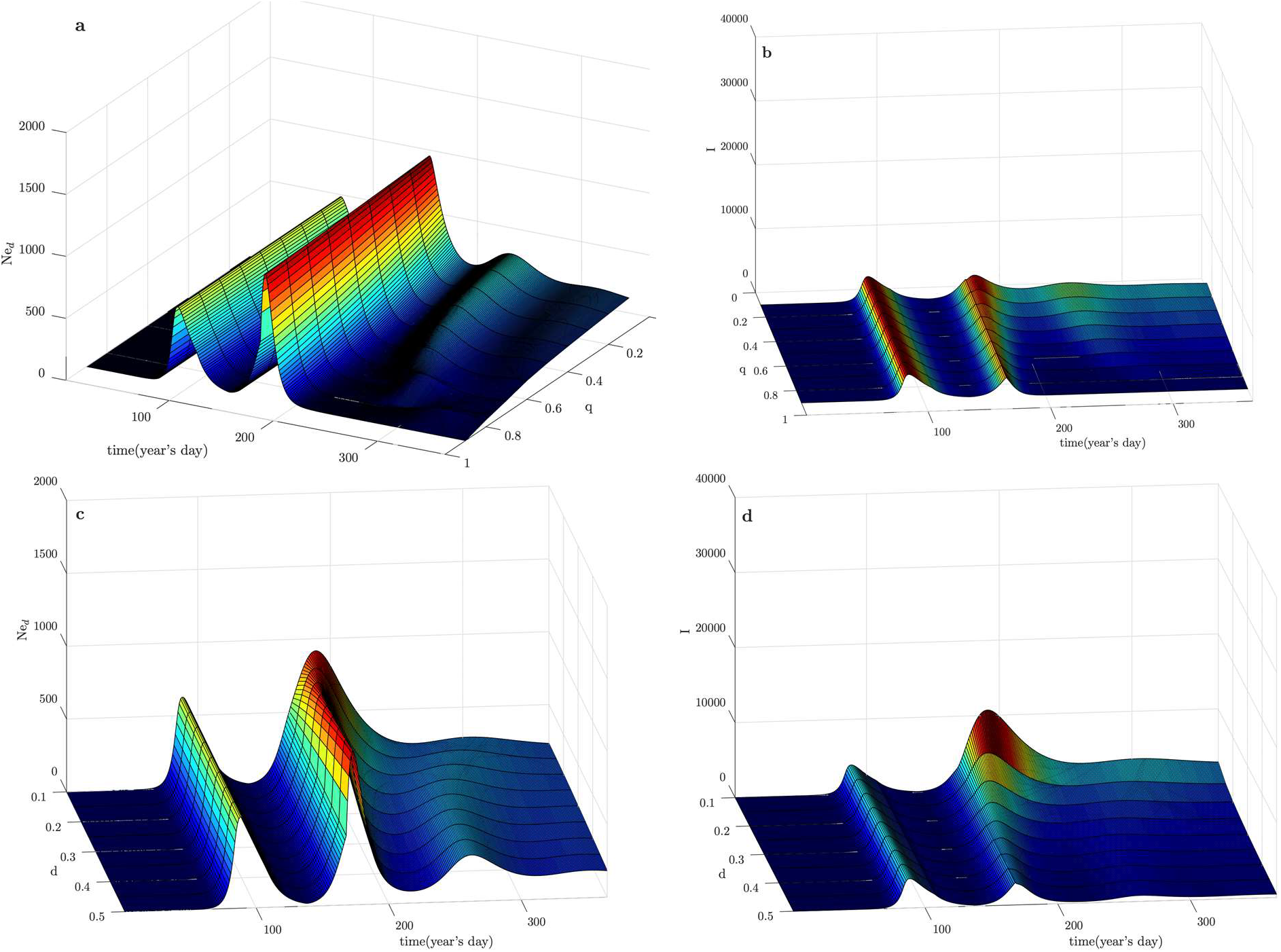
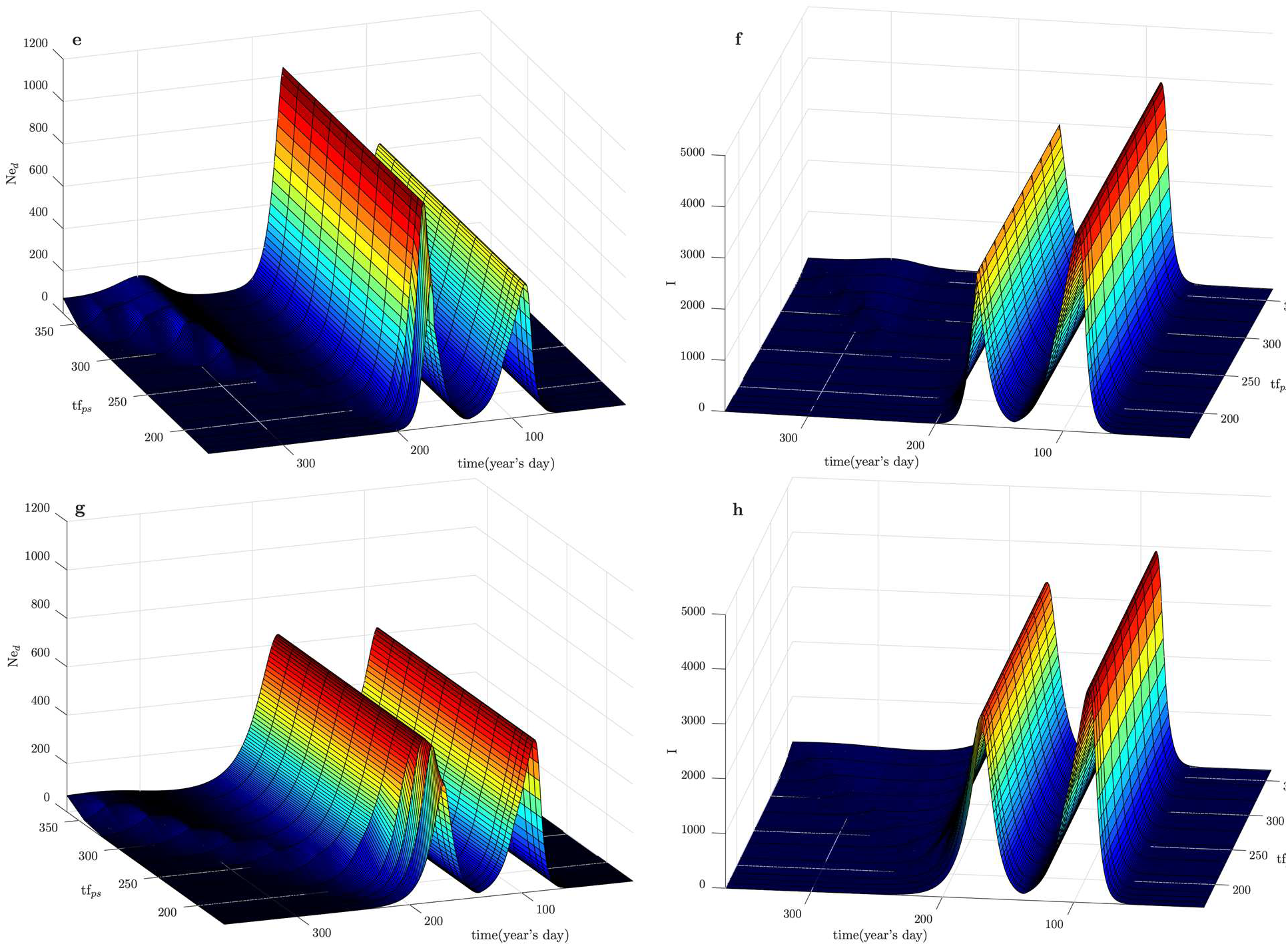
Model simulation for **IL** based on ***Ne*_d_** (first column) and ***I*** (second column) time evolutions. Parameters change only after day 167. (**a**) and (**b**): ***q*** is changed for ***d*** = 0.5. (**c**) and (**d**): ***d*** is changed for ***q*** = 0.5. (**e**) and (**f**): Different ***PS***-flow stopping moments for ***d*** = 0.5 and ***q*** = 0.75. (**g**) and (**h**): Different ***PS***-flow stopping moments for ***d*** = 0.1 and ***q*** = 0.9.

Figure 6a-b show the effect of varying ***q*** for a fixed ***d*** and Fig 6c-d the effects of varying ***d*** for a fixed ***q***. The fixed values of ***q*** and ***d*** were both set to 0.5. When such conditions are applied, it is possible to observe that it is much worse to have a smaller value of ***d*** than to have a smaller value of ***q***. However, ***q*** should not be small if ***d*** is 0.1, which is the case for the majority of the analyzed countries. If this happens, a small ***q*** can have a catastrophic effect (Supplementary Figure S4).

Figure 6e-h explores the variation of the moment when the ***PS***-flow is stopped. Again, a further confirmation that increasing ***d*** is a better option than increasing ***q*** as well as the fact that stopping the leakage as early as possible is the only way of fully stopping the epidemic

Taken together, these simulations indicate that massive testing and contact tracing are fundamental in containing the disease and that the action taken must never rely solely on ***q***.

## Discussion

The method presented in this work was successfully applied and enabled the analysis of the data from the COVID-19 pandemic taking advantage of an open-access online fitting plateform. By explaining the data from the past it was possible to obtain more robust projections for the future and evaluate the actions that can be taken to prevent unsustainable spreading of the disease.

The model was successfully fitted to a large number of data compartments, in eleven different countries with large geographical distribution and diversity of dynamics and were able to explain the complex dynamics of disease spreading, including second waves and identification of the date of important past behavioral changes. It is important to point out that the selection of the countries relied on their consistent and systematic reporting of data. For not correctly reporting the number of daily recovered, some countries could not be a part of this analysis. However, there is no reason to believe that the model is not able to accurately fit any good quality data including systematic counts of infected, recovered, and dead, provided by any country.

Interestingly, the flow to and from the ***P*** compartment can be discussed under the perspective of risk perception levels. If the risk perception is high, people will naturally flow to ***P*** even without government-imposed measures. This is most likely what happened in the early days of the pandemic, when a reduction in mobility was observed prior to imposed lockdowns. Conversely, if risk perception lowers, the flow to ***P*** will decrease while the flow to ***S*** is more likely to increase and lead to secondary outbursts. This suggests that all mitigation policies should include clear communication strategies to assure population compliance. Neglecting the role of communication or providing contradictory information can severely render the efforts of testing and confinement, especially in democratic countries.

The method presented in this work enables public health authorities to quantify the effect of alternatives to lockdown, namely massive testing and self-protection measures. The former decreases the number of silent spreaders, altough it may initially produce an increase in the detected cases, while the latter decreases the ***PS***-leakage and increases the value of ***q*** (fraction of exposed that end up not spreading the disease). Furthermore, it was demonstrated that policies relying merely on increasing the value of ***q*** have a very low chance of producing a positive result, since the number of complying people needs to be unrealistically high.

Overall, as both the model and the system to test different parameters is open and freely available, we hope to encourage other colleagues to analyze further countries, more complex models, and parameter space(s), projecting potential future scenarios and, by doing so, understand the actions that can be taken to effectively prevent an endemic characteristic of the disease in the future.

We expect that the knowledge and the model provided in this work can be of use to governmental authorities for mitigating disease spread while waiting for effective vaccines or treatments.

## Methods

### Data

We have applied the PSEIRD(S) model to systematically analyze the data from eleven countries: Italian Republic, **IT**, Federal Republic of Germany, **DE**, Canada, **CA**, Islamic Republic of Iran, **IR**, Japan, **JP**, Swiss Confederation, **CH**, Israel, **IL**, Republic of Austria, **AT**, Republic of Korea, **KR**, Iceland, **IS**, and New Zealand, **NZ**. These countries were selected following four criteria: geographic, cultural and dynamical variety, having reached the (first wave) pandemic peak and having provided systematic data on the number of infected, recovered, and deceased.

The evolution of the pandemic, for the analyzed countries, includes daily reports of the number of new cases and the cumulative number of deaths and recovered that were collected from the Johns Hopkins GitHub time series data base^19^. These represent three independent data sets from which two additional sets can be obtained: the total number of cases and the active infected (detected) cases.

### Model fitting and simulations

A total of five data sets could be simultaneously analyzed providing the estimation for the parameters described in Supplementary Table S1. Following this analysis, it was possible to simulate the remaining compartments based on the optimized model parameters.

The model fits were performed using an open-access user-friendly online platform, maintained by the authors – *fitteia®*^20^. – that employs the non-linear least-squares minimization method and is accessible at http://fitteia.org.

As epidemiological models usually require the resolution of differential equations, the Runge-Kutta method integrated in fitteia*®* was used in the minimization process. After carefully setting the initial population values for each compartment and initial parameter values, a Runge-Kutta iteration with a time resolution “*h*” (always fixed to 0.01) was applied. Then, the solutions for each equation were compared with the existing data in a loop until a global minimum in the model parameters space is obtained. *fitteia®* uses the powerful and efficient minimization process provided by the numerical routine MINUIT from the CERN library^21^.

## Data Availability

All data is available in the main text, the supplementary materials, or is from public repositories (https://github.com/CSSEGISandData/COVID-19). All code and fitting results are available online at https://github.com/fitteia/COFIT.

https://github.com/fitteia/COFIT

## Supplementary materials

Are available at https://github.com/fitteia/COFIT.

## Acknowledgments

P.J.S wishes to thank Luis Gonçalves for fruitful discussions.

## Author contributions

Conceptualization: all authors; Tool development: PJS; Model fitting and simulations: P.J.S, M.J.B, A.K. Paper writing: all authors.

## Data and materials availability

All data is available in the main text, the supplementary materials, or is from public repositories. All code and fitting results are available online at https://github.com/fitteia/COFIT.

